# An assessment of demographics, clinical features and risk factors in patients with chronic respiratory disorders in two Latin American countries

**DOI:** 10.1101/2025.03.13.25323731

**Authors:** Albert Farrugia, Claudia Jiménez, Rafael Salazar, Eva Maroto, Andrea Gonzales, Rosario D’Imperio, Shane Fitch

## Abstract

Lovexair is a non-profit organization dedicated to the care, support and guidance of people with respiratory pathologies and to the support of healthcare professionals. Lovexair has developed the **HappyAir Community**, an ecosystem of clinical, educational and social resources for comprehensive patient care, well-being and a better quality of life. We report on the outcomes of events held in two Latin American countries (Colombia and Mexico) in late 2023, launching the HappyAir project in these communities through assessing the demographics, clinical and risk factors and outcomes of respiratory diseases on patients affected by these conditions. A total of 103 patients with respiratory problems were assessed, using spirometry, the exhaled nitric oxide test (FeNO), the COPD Assessment Test (CAT) and the Asthma Control Test. Only 22% of the study population showed abnormal spirometry, despite a prevalence of asthma and COPD of 39%. Conversely, the FeNO test was abnormal to the same extent as asthma was prevalent. This population also showed an association between BMI and dyspnoea. The subjects showed a high level of awareness on the effect of environmental pollution on their respiratory problems. The experience and results of this exercise will be used to inform and guide clinical and social interventions aiming to improve the prospects and quality of life in patients with chronic respiratory disorders. They should assist in identifying and addressing diagnostic and treatment gaps in communities suffering from respiratory illness.

## Introduction

Lovexair (https://www.lovexair.com/en/lovexair-foundation/) is a non-profit organization dedicated to the care, support and guidance of people with respiratory pathologies and to the support of healthcare professionals. Lovexair’s vision is to offer respite to people affected by chronic respiratory diseases, their families and caregivers, through raising awareness in society about respiratory diseases and their impact. Lovexair seeks to provide innovative solutions and tools in digital health to avoid these diseases, improve the quality of life and well-being of the affected person in their family environment, and to support health professionals and public and private health services. Lovexair has developed the **HappyAir Community**, an ecosystem of clinical, educational and social resources for comprehensive patient care, well-being and a better quality of life. We report on the outcomes of events held in two Latin American countries (Colombia and Mexico) in late 2023, launching the HappyAir project in these communities through assessing the demographics, clinical and risk factors and outcomes of respiratory diseases on patients affected by these conditions.

## Methods

Patients with respiratory disorders were invited to participate in two workshops, one in Colombia, one in Mexico. Thirteen health care professionals trained in assessing respiratory illnesses through spirometry and other respiratory function tests described in the Results. During the workshops, these health care professionals conducted interviews with patients to collect information about their respiratory diseases, comorbidities, symptoms, risk factors (smoking, internal and external contamination, family history) exacerbations, triggers, treatment and adherence, pursuing an active life, and daily activities. Patient quality of life, the patients’ overall state of health and their respiratory function were assessed and recorded. A total of 103 patients with respiratory problems were assessed.

During the workshops, health care All patients received a clinical report and educational contents about lung health. After the workshops, patients were invited to attend webinars about their diseases.

Health care professionals did little meetings with patients to teach us different aspects about their diseases (inhalers, exercises, how to manage their symptoms, diaphragmatic breathing)

After the workshops, patients were invited to attend to different webinars about their diseases.

## Results

The patient demographic did not vary significantly between the two countries and is summarised as follows:

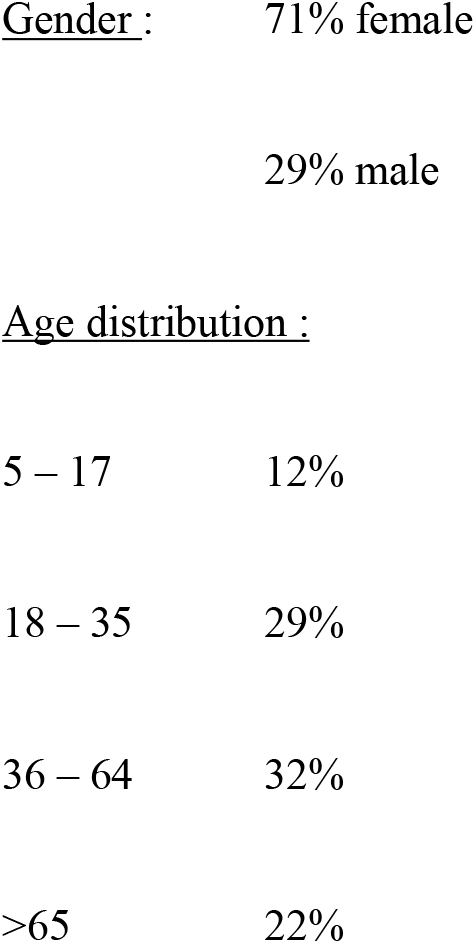

The spectrum of previously diagnosed disorders is shown on **Fig 1**. Further assessment carried out over the course of the HappyAir events included spirometry (1), the exhaled nitric oxide test (FeNO) (2), the COPD Assessment Test (CAT) (3) and the Asthma Control Test (ACT) (4). In addition, a subset of patients from Colombia were assessed for their respiratory health status relative to body mass index (BMI) and for the effect of their respiratory health status on their emotional health. In addition, the degree of dyspnoea was assessed using the MRC scale (5). The results of these tests are shown in **Table 1**.

**Table 1.** Test results in the study population.

| Patients | % |
| --- | --- |
| Abnormal spirometry | 22 |
| Abnormal FeNO | 29 |
| Undiagnosed patients who reported dyspnoea | 69 |
| High BMI indicating overweight and obese | 55 |
| High BMI reporting dyspnoea | 62 |
| Uncontrolled asthma (on basis of ACT) | 14 |
| Severe/very severe COPD (on basis of CAT) | 50 |
| Reporting high impact from environmental pollution | 43 |
| Considering impact of pollution significant | 37 |
| Ex/still/passive smokers | 24 |
| Respiratory illness affects their work | 20 |
| Severe emotional issues | 44 |

**Figure 1.**
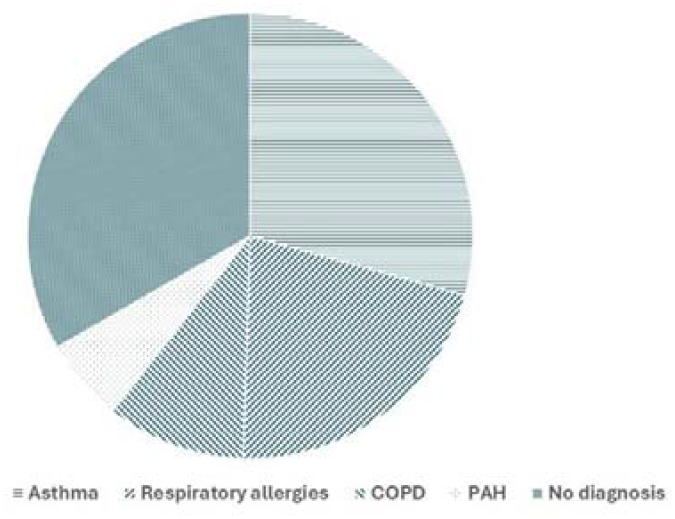
Previously diagnosed disorders in the study population.

## Discussion

These results support other reports indicating the occasional limitation of spirometry in supporting a diagnosis of chronic respiratory disorders such as COPD and asthma (comprising 39% of the studied population with only 22% showing abnormal spirometry (6,7). Conversely, the FeNO test was abnormal to the same extent as asthma was prevalent – 29%, confirming the utility of this test in asthma diagnosis and management (8). Enhancing diagnostic capacity, through these and other tests, is clearly important in the geographies studied, given the high level of patients with undiagnosed disease. This population also showed an association between BMI and dyspnoea, as has been shown in other patient groups (9). Importantly, the study subjects showed a high level of awareness on the effect of environmental pollution on their respiratory problems, an aspect confirmed in other studies (10).

The outcomes from these and other HappyAir activities will be used to inform and guide clinical and social interventions aiming to improve the prospects and quality of life in Lovexair’s target constituency. They should assist in identifying and addressing diagnostic and treatment gaps in communities suffering from respiratory illness in countries which are “under the radar” of the therapeutic landscape for these widespread health problems.

## Data Availability

All data produced in the present study are available upon reasonable request to the authors

## References

1. Ponce MC, Sankari A, Sharma S. Pulmonary Function Tests. In: StatPearls [Internet]. Treasure Island (FL): StatPearls Publishing; 2024 [cited 2024 Feb 6]. Available from: http://www.ncbi.nlm.nih.gov/books/NBK482339/

2. Harnan SE, Tappenden P, Essat M, Gomersall T, Minton J, Wong R, et al. Measurement of exhaled nitric oxide concentration in asthma: a systematic review and economic evaluation of NIOX MINO, NIOX VERO and NObreath. NIHR Journals Library; 2015.

3. Lin JS, Webber EM, Thomas RG. Introduction to Screening for Chronic Obstructive Pulmonary Disease. In: Screening for Chronic Obstructive Pulmonary Disease: A Targeted Evidence Update for the US Preventive Services Task Force [Internet] [Internet]. Agency for Healthcare Research and Quality (US); 2022 [cited 2024 Feb 6]. Available from: https://www.ncbi.nlm.nih.gov/books/NBK580644/

4. Przybyszowski M, Stachura T, Szafraniec K, Sladek K, Bochenek G. The influence of self-assessment of asthma control on the Asthma Control Test outcome. J Asthma Off J Assoc Care Asthma. 2021 Apr;58(4):537–46.

5. Fletcher CM, Elmes PC, Fairbairn AS, Wood CH. The significance of respiratory symptoms and the diagnosis of chronic bronchitis in a working population. Br Med J. 1959 Aug 29;2(5147):257–66.

6. Arne M, Lisspers K, Ställberg B, Boman G, Hedenström H, Janson C, et al. How often is diagnosis of COPD confirmed with spirometry? Respir Med. 2010 Apr 1;104(4):550–6.

7. Fortis S, Corazalla EO, Kim HJ. Does normal spirometry rule out an obstructive or restrictive ventilatory defect? Respir Investig. 2017 Jan 1;55(1):55–7.

8. Karrasch S, Linde K, Rücker G, Sommer H, Karsch-Völk M, Kleijnen J, et al. Accuracy of FENO for diagnosing asthma: a systematic review. Thorax. 2017 Feb;72(2):109–16.

9. Wang Z, Zhou X, Deng M, Yin Y, Li Y, Zhang Q, et al. Clinical impacts of sarcopenic obesity on chronic obstructive pulmonary disease: a cross-sectional study. BMC Pulm Med. 2023 Oct 18;23(1):394.

10. Yan P, Liu P, Lin R, Xiao K, Xie S, Wang K, et al. Effect of ambient air quality on exacerbation of COPD in patients and its potential mechanism. Int J Chron Obstruct Pulmon Dis. 2019;14:1517–26.

